# Modeling infection dynamics and mitigation strategies to support K-6 in-person instruction during the COVID-19 pandemic

**DOI:** 10.1101/2021.02.27.21252535

**Authors:** Douglas E. Morrison, Roch Nianogo, Vladimir Manuel, Onyebuchi A. Arah, Nathaniel Anderson, Tony Kuo, Moira Inkelas

## Abstract

**Objective:** To support safer in-person K-6 instruction during the coronavirus disease 2019 (COVID- 19) pandemic by providing public health authorities and school districts with a practical model of transmission dynamics and mitigation strategies.

**Methods:** We developed an agent-based model of infection dynamics and preventive mitigation strategies such as distancing, health behaviors, surveillance and symptomatic testing, daily symptom and exposure screening, quarantine policies, and vaccination. The model parameters can be updated as the science evolves and are adjustable via an online user interface, enabling users to explore the effects of interventions on outcomes of interest to states and localities, under a variety of plausible epidemiological and policy assumptions.

**Results:** Under default assumptions, secondary infection rates and school attendance are substantially affected by surveillance testing protocols, vaccination rates, class sizes, and effectiveness of safety education.

**Conclusions:** Our model helps policymakers consider how mitigation options and the dynamics of school infection risks affect outcomes of interest. The model’s parameters can be immediately updated in response to changes in epidemiological conditions, science of COVID-19 transmission dynamics, testing and vaccination resources, and reliability of mitigation strategies.

## Introduction

School closures due to the coronavirus disease 2019 (COVID-19) pandemic harm children’s educational progress, psychosocial development, and mental and physical health.^1–4^ School closures also have cascading economic and social impacts on families, workplaces, and workforce participation.^5^ Since COVID-19 burden is often greater in socioeconomically disadvantaged communities, in-person instruction has been unavailable or inconsistent for the students who may need it the most, thereby increasing educational disparities for children in lower income families, communities of color, and/or households that include essential workers, contain multiple generations, experience crowded housing, and with members who have high-risk chronic conditions (e.g., hypertension, diabetes, obesity).^6,7^

Yet uncertainty surrounding infectiousness and management of pre- and asymptomatic populations has slowed school re-opening in the U.S. Resuming in-person instruction during the pandemic requires bringing susceptible children and adults into a congregate setting for extended periods of time with daily return into their communities. Recent studies suggest that secondary infection risk in schools is low when basic precautions are followed.^8–10^ Yet safety concerns remain for local governments, public health authorities, school district administrators, labor unions, parents, and students. Public health authorities and school districts in communities with higher COVID-19 prevalence have concerns about the possibility of secondary infection associated with in-person instruction. Initially, school re-opening was frequently tied to regional COVID-19 case rates and testing positivity thresholds; more recently, the Centers for Disease Control and Prevention (CDC) is stressing the enabling impact of mitigation strategies even when COVID-19 case rates are high.

To understand and act upon risk in a dynamic environment, it is essential that public health authorities and districts understand the levers that are available to them. There is a paucity of practical models that inform planning for safer in-person instruction in K-6 settings. Moreover, most models to-date are predictive and often a black box, designed to provide a single best answer, rather than illustrative with the goal of promoting understanding about levers of influence, which is vital for population health decision-making.^11^

To help address this gap in public health practice, we designed a stochastic, agent-based model of resumed in-person instruction that includes representations of a variety of intervention levers, including screening for symptoms and exposures, biological testing, education to reduce risky behaviors, and vaccination. Recognizing that the science of COVID-19 is rapidly evolving, we sought to create a model with parameters that end users could easily adjust as more information emerges regarding transmission dynamics and the impact of mitigation strategies. Additionally, we sought flexibility to accommodate characteristics specific school communities and their local environments. We developed this model while collaborating as a university science partner with a large urban school district to consider what would be necessary for safer resumption of this much needed face-to-face learning. We assess key outcomes of interest to school district stakeholders, and we provide an online user interface to the model as a practical tool to empower school districts and their implementation partners to explore how various combinations and variations of strategy components affect health and learning outcomes within different underlying epidemiological conditions that could arise in the real world. This user interface can be accessed at https://agent-based-models.shinyapps.io/RegionalCOVIDSchoolSimulation/.

## Methods

### Model design and scope

Here we describe the model in general terms; a detailed description of the implemented model is provided as Supporting Information. The R code implementing the model is available on request from the corresponding author.^12^

#### Agents

The model contains two types of simulated individuals (“agents”): students and their associated household adults (two per student). We chose to include these agent types as they form the largest proportion of the school community. The model could be extended to include teachers and other school personnel as additional agents who interact with students and with one other. Each student is assigned to a particular school and classroom and has a number of “close classmates” within their class; close classmates have higher risks of transmission than other classmates.

#### Sources of infection

Infections in the model come from three sources: infectious classmates at school, infectious family members at home, or exogenous exposures (a catch-all category for in-person work, non-distanced socializing, grocery shopping, or any other exposures outside of schools and households).

#### Infection progression

Agents follow a susceptible-infected-recovered (SIR) framework in which they are initially either susceptible to infection, infected, or vaccinated or recovered (i.e. immune).^13^ Susceptible individuals can become infected over the course of the simulation. Infected individuals then progress through a series of infection states. They first enter a latent period during which they are not yet infectious or symptomatic. Next, they become infectious but still presymptomatic. Infectious presymptomatic individuals can become symptomatic or remain asymptomatic. Eventually, infected individuals recover and become immune.

#### Interventions

The model includes representations of a number of possible program components for resuming and maintaining in-person instruction. Not all of these components may be implemented in some school districts, so the model has options for some of these components to be deactivated. For example, surveillance testing of nonsymptomatic individuals can be eliminated by setting the “testing fraction” for surveillance testing to 0%. Thus, model users can modify parameters to exclude or alter some of the mitigation strategies to represent scenarios relevant to their environment.

One component is a daily health screening (“attestation”) system through which school community member are prompted to self-report suspected COVID-19 exposures and symptoms. Daily health screenings may reduce the rates of infectious individuals coming onto campus; individuals reporting symptoms or suspected COVID-19 exposures could be diverted into quarantine protocols or receive other triage and follow-up. Such screenings have been implemented in some workplaces, universities, and K-12 systems.^14^

Another component is education of the school community members about safe behaviors that reduce their exposure to COVID-19. In addition to influencing behavior, such engagement of students and their families may change the likelihood that individuals accurately report potential exposures and symptoms on the daily screen. The model includes a representation of this possible mechanism.

Testing for COVID-19 infection is another possible component. Tests can be performed reactively when a school community member (student or parent/guardian) reports COVID-19 symptoms or suspects they have been exposed. Periodic surveillance testing can also be performed proactively on random samples of nonsymptomatic individuals to identify presymptomatic and asymptomatic cases. Both types of testing are represented in the model. The model can represent tests with different accuracy characteristics (specificity, sensitivity as a function of elapsed time since infection). In the analyses below, we assumed accuracy characteristics similar to PCR testing, but these parameters and testing frequency could be changed to represent antigen testing.^15^

Other policies that can influence infection risks at school include defining and maintaining small groups in close proximity (e.g., classrooms, lunch groups) as well as using masks, physical space dividers, and other forms of physical barriers.^16^

### Model outcomes

We report the following model outcomes after two months of simulated full-time in-person school:

1. The cumulative percentage of enrolled students infected with COVID-19 since baseline.
2. The cumulative percentage of enrolled students infected with COVID-19 while at school.
3. The cumulative number of school days missed per student.
4. The percentage of schools with no in-school transmissions.
5. The percentage of schools with no detected infection clusters.

Other measured outcomes, such as infection rates among household adults, are not reported here in the interest of brevity but are provided in the online interface.

For each experimental scenario considered below, we simulated 1000 schools in a single run of the model using the corresponding set of input parameter values. We calculated the five outcomes listed above for each school, and then combined results across the simulated schools to estimate outcome distribution summary statistics. For the numeric outcomes (#1-3), we report the means across the 1000 simulated schools, as well as the 2.5% and 97.5% percentiles of these outcomes as 95% prediction intervals. For the binary outcomes (#4-5) we report the event rates as percentages (prediction intervals are not applicable for binary outcomes).

### Validation and calibration

Validation tests confirmed that the agent-initializing function produced the specified initial rates of current infection, prior infection, and COVID safety education characteristics among students at baseline for the default input parameter values. In the absence of data on school re-openings at the time of publication, it was not feasible to calibrate this model. As data emerge from school districts across the U.S., it will be possible to quantitatively calibrate and validate the model in future work.

For this paper, we assigned default parameter values based on the existing literature where possible and considered likely values for variables with considerable uncertainty (details in Supporting Information). To use the model to inform planning, policymakers should choose parameter values that reflect current conditions in their schools.

### Example experiments

To demonstrate how the model can be used to explore the effects of interventions, we tested the effects of changes in four parameters that could be affected by school policies: class size, frequency of surveillance testing, fraction of students tested in each surveillance sample (“sampling fraction”), and proportion of household adults vaccinated (Tables 1-3). We started with the default parameter values and varied these four parameters to determine how the outcomes changed in response. We also considered a set of four scenarios examining interactions between surveillance testing and community education (Table 4).

**Table 1:** Mean-average outcomes after two months of in-person instruction (with 2.5% and 97.5% percentiles for numeric outcomes) for 1000 simulated schools, by class size, with all other parameters set to default values.

| Number of students per class | % of enrolled students infected since baseline (cumulative) | % of enrolled students infected from school (cumulative) | # schooldays quarantined per student (cumulative) | % of schools with no on-campus transmissions so far | % of schools with no detected infection clusters so far |
| --- | --- | --- | --- | --- | --- |
| 10 | 4.49 (2.62, 6.67) | 0.04 (0.00, 0.24) | 1.20 (0.92, 1.51) | 85.9 | 99.5 |
| 15 (default) | 4.46 (2.62, 6.67) | 0.05 (0.00, 0.48) | 1.20 (0.93, 1.55) | 82.0 | 98.9 |
| 20 | 4.47 (2.62, 6.67) | 0.06 (0.00, 0.48) | 1.21 (0.90, 1.58) | 80.2 | 97.9 |
| 30 | 4.52 (2.62, 6.91) | 0.09 (0.00, 0.48) | 1.23 (0.91, 1.79) | 68.3 | 95.1 |

**Table 4:** Scenarios assessing the effects and interactions of surveillance testing and educational outreach to families; mean-average outcomes (with 2.5% and 97.5% percentiles for numeric outcomes) for 1000 simulated schools, with all other parameters set to default values.

| Scenario Description | Surveillance testing frequency | Probability of outreach receptiveness | Improvements in attestation accuracy and exogenous risk from COVID safety education | % of enrolled students infected since baseline (cumulative) | % of enrolled students infected from school (cumulative) | # schooldays quarantined per student (cumulative) | % of schools with no on-campus transmissions so far | % of schools with no detected infection clusters so far |
| --- | --- | --- | --- | --- | --- | --- | --- | --- |
| Default parameter values | 25% every Monday | 50% | 10% | 4.46 (2.62, 6.67) | 0.05 (0.00, 0.48) | 1.20 (0.93, 1.55) | 82.0 | 98.9 |
| No testing or outreach | None | 0% | 50% | 4.39 (2.62, 6.43) | 0.06 (0.00, 0.48) | 1.15 (0.85, 1.46) | 78.7 | 99.4 |
| Education only | None | 100% | 50% | 4.10 (2.38, 6.19) | 0.05 (0.00, 0.48) | 1.08 (0.81, 1.38) | 82.3 | 99.7 |
| Testing only | 100% every Monday | 0% | 50% | 4.31 (2.38, 6.43) | 0.03 (0.00, 0.24) | 1.25 (0.94, 1.62) | 86.9 | 96.5 |
| Testing + education | 100% every Monday | 100% | 50% | 4.07 (2.38, 5.95) | 0.03 (0.00, 0.24) | 1.16 (0.88, 1.50) | 89.0 | 97.5 |

### Sensitivity analyses

In sensitivity analyses, we tested the effects of additional parameters: number of non-socially distanced classmates per student, test sensitivity and specificity, transmission risk from infectious students to non-socially-distanced classmates, exogenous infection risk, attestation sensitivity and specificity prior to COVID-19 safety education outreach, receptiveness to COVID-19 safety education outreach, and effects of COVID safety education on attestation accuracy and exogenous risk (Supporting Information). We created tornado plots as a simple, interpretable display; these plots assess the relative importance of these variables with respect to each outcome.^17^

## Results

### Example experiments

In 1000 schools simulated for two months of in-person instruction using the default parameter values except for class size, we found that smaller class sizes resulted in fewer students infected in-school, more schools remaining transmission-free, and fewer schooldays missed (Table 1). With classes of 30 students each, an average of 4.52% of students became infected, 0.04% were infected in-school, 1.23 schooldays were missed per student, and 68.3% of schools remained transmission-free. With 10 students per class, these outcomes improved to 4.49% infected overall, 0.04% infected in-school, 1.20 schooldays missed per student, and 85.9% of schools remaining infection-free.

More surveillance testing resulted in lower transmission rates, but more schooldays missed (Table 2). With no surveillance testing, 4.48% of students became infected after baseline, 0.05% were infected at school, 1.18 schooldays were missed per student, and 81.4% of schools had no on-campus transmissions. Weekly surveillance testing with 25% sampling fractions did not change these outcomes substantially. Daily testing with 25% sampling produced noticeable improvements in infection rates: 4.42% of students became infected overall, 0.04% were infected in school, and 84.5% of schools remained transmission-free; however, average schooldays missed increased to 1.26 days per student. Finally, daily testing of all students produced larger improvements in secondary infection rates: 4.43% of students were infected overall, 0.03% in school, and 88.2% of schools stayed transmission-free; however, schooldays missed rose to 1.57 days per student. The percentage of schools with no detected clusters had an opposite trend to the percentage of schools with no actual transmissions: with more surveillance testing of asymptomatic students, more schools had clusters detected.

**Table 2:** Mean-average outcomes after two months of in-person instruction (with 2.5% and 97.5% percentiles for numeric outcomes) for 1000 simulated schools, by surveillance testing frequency and sampling fraction, with all other parameters set to default values.

| Sampling fraction | Surveillance testing schedule | % of enrolled students infected since baseline (cumulative) | % of enrolled students infected from school (cumulative) | # schooldays quarantined per student (cumulative) | % of schools with no on-campus transmissions so far | % of schools with no detected infection clusters so far |
| --- | --- | --- | --- | --- | --- | --- |
| 0% | No surveillance testing | 4.48 (2.62, 6.67) | 0.05 (0.00, 0.48) | 1.18 (0.89, 1.51) | 81.4 | 99.5 |
| 25% | Once a week (M) (*default*) | 4.46 (2.62, 6.67) | 0.05 (0.00, 0.48) | 1.20 (0.93, 1.55) | 82.0 | 98.9 |
|  | Twice a week (M/Th) | 4.49 (2.62, 6.67) | 0.05 (0.00, 0.24) | 1.23 (0.94, 1.58) | 81.7 | 97.5 |
|  | 3x a week (MWF) | 4.50 (2.62, 6.43) | 0.05 (0.00, 0.24) | 1.26 (0.96, 1.60) | 80.3 | 95.9 |
|  | Every weekday (M-F) | 4.45 (2.62, 6.43) | 0.04 (0.00, 0.24) | 1.29 (0.98, 1.66) | 83.5 | 94.9 |
| 100% | Once a week (M) | 4.42 (2.86, 6.20) | 0.04 (0.00, 0.24) | 1.26 (0.98, 1.63) | 84.5 | 95.5 |
|  | Twice a week (M/Th) | 4.47 (2.62, 6.43) | 0.04 (0.00, 0.24) | 1.35 (1.02, 1.75) | 86.5 | 90.4 |
|  | 3x a week (MWF) | 4.44 (2.62, 6.43) | 0.03 (0.00, 0.24) | 1.43 (1.07, 1.86) | 88.2 | 83.5 |
|  | Every weekday (M-F) | 4.43 (2.62, 6.43) | 0.03 (0.00, 0.24) | 1.57 (1.19, 2.08) | 88.2 | 73.0 |

Higher levels of vaccination among household adults resulted in fewer infections overall and fewer schooldays missed, but no improvements in on-campus infections (Table 3). With no vaccinations, 4.46% of students became infected, 0.05% were infected while at school, 1.20 days were missed, and 82.0%% of schools had no on-campus transmissions. With 75% of the adults vaccinated, only 3.90% of students became infected and 0.79 schooldays were missed per student; 0.06% of students were infected on campus, and 80.1% of schools had no on-campus transmissions. This is expected given that the default parameter value for the risk of parent-to-child transmission is low (16% per infection; details in Supporting Information).

**Table 3:** Mean-average outcomes (with 2.5% and 97.5% percentiles for numeric outcomes) for 1000 simulated schools, by adult vaccination rate, with all other parameters set to default values.

| Percentage of household adults vaccinated | % of enrolled students infected since baseline (cumulative) | % of enrolled students infected from school (cumulative) | # schooldays quarantined per student (cumulative) | % of schools with no on-campus transmissions so far | % of schools with no detected infection clusters so far |
| --- | --- | --- | --- | --- | --- |
| 0% (*default*) | 4.46 (2.62, 6.67) | 0.05 (0.00, 0.48) | 1.20 (0.93, 1.55) | 82.0 | 98.9 |
| 25% | 4.15 (2.38, 6.19) | 0.05 (0.00, 0.24) | 1.06 (0.79, 1.35) | 81.2 | 99.1 |
| 50% | 3.90 (2.14, 5.71) | 0.07 (0.00, 0.48) | 0.92 (0.68, 1.18) | 76.5 | 99.0 |
| 75% | 3.62 (1.91, 5.48) | 0.06 (0.00, 0.48) | 0.79 (0.59, 1.03) | 80.1 | 98.9 |

In the scenarios examining interactions between surveillance testing and school community education, we found that the “Education only”, “Testing only”, and “Testing + Education” scenarios all resulted in lower infection rates than the “No testing or education” scenario (Table 4). “Testing only” had better in-school infection rates than “Education only” but worse overall infection rates and average attendance rates. “Testing + Education” had better infection rates than either strategy alone and a better average attendance rate than “Testing alone”.

### Sensitivity analyses

Detailed sensitivity analysis results are provided in the Supporting Information. Briefly: starting from our default assumptions, we found that in-school infections were most affected by changes in risk per infectious classmate, exogenous infection risk, exposure and symptoms screening sensitivity and specificity, likelihood of symptoms if infected, class size, number of non-distanced classmates in school, and biological test specificity, while attendance rates were most affected by changes in symptoms/exposure screening specificity, exogenous infection risk, biological test specificity, and vaccination rate.

## Discussion

Public health credibility has been vital in the COVID-19 pandemic, so models must reflect the latest science to be informative and trusted. This means building adaptability into models intended to guide planning. Our model supports public health practice with a practical, transparent, and evidence-based representation of known mechanisms for transmission and mitigation and their interdependencies. Critically, the model has explicit assumptions and is adaptable, which is essential within a rapidly evolving scientific context. Effectively communicating the latest scientific thinking to key decision makers has been a challenge throughout the current pandemic and will likely be for future pandemics as well. Models have great potential value in surfacing inputs, outputs, and decision-points even when parameters are unknown or emergent. Transparent assumptions, and the ability to modify them as COVID-19 science evolves, offers decision-makers a way of understanding the potential impact of their decisions under complex conditions. This model will have increasing value for predicting the impact of different decisions as there is greater certainty from science regarding specific mitigation efforts (e.g., effectiveness of facial coverings) as well as infection and transmission dynamics.

With equity of school re-opening in mind, considering mitigation strategies under different conditions is essential for modeling. Using parameters that reflect our current best knowledge about COVID-19, the model suggests that no single program element or condition ensures safety and also shows that schools could employ multiple elements that work together. Our model reflects what recent studies suggest; even without COVID-19 testing, good on-campus infection control has been shown to reduce on-campus transmission, and high community prevalence does not necessarily lead to significant secondary infection.^10^

Notably, the model illustrates the value of school districts measuring not just adoption of policy but the implementation quality of their mitigation strategies. When using plausible values for key parameters, key outcomes in our model are influenced by the sensitivity and specificity of mitigation efforts, such as the extent to which self-reporting of symptoms and exposures reflect actual risk (infectiousness). Given the presymptomatic and asymptomatic features of COVID-19, particularly in children, the model helps decision-makers appreciate the impact of accurate reporting of symptoms and exposures. This shows the potential value of assessing accuracy within a school system and iteratively improving it through education, effective design of the health check questions or online application/portal, or other means. Notably, while our model includes both symptoms and possible exposure in the daily health screening, current CDC guidance notes limitations of using symptoms in a daily health screening among K-12 students.^14^

Modeling possible outcomes under different scenarios helps to show how local conditions and program choices affect outcomes that stakeholders care about. As the pandemic progresses, more decision-makers are involved, and each locality has a unique set of political, resource, and behavioral constraints. This model enables localities to see the implications of these constraints and to identify the combination of elements that could work given their constraints. The results flowing from these models can in turn be used to empower action by providing stakeholders with a simple data display and easy interface for parameter estimation, which can in turn inform collective actions may be needed to achieve specific goal targets that they establish for their system. This includes governance, public health, school district administrators, labor unions, parents, and students as well as the broader communities. Models such as this one are useful for considering equity implications in areas with significant regional variation in factors ranging from COVID-19 prevalence to the knowledge and ability to be forthcoming in reporting possible exposures and symptoms.

A critical feature of this model is that its parameters can be adapted based on the evolution of COVID- 19 science and the technology of testing and vaccination and adjusted as school districts generate data in their own environments. Just as it has been well-established that a “one-size fits all” approach is insufficient for testing, treatment, and vaccination, social processes like school transmission benefit from locally tailored approaches.^18–20^ Models that account for differences in underlying populations and social dynamics are a valuable tool for adopting a “precision medicine” approach to population health.^18^

Finally, the model structure and dynamics are not limited to COVID-19. Experience with the COVID- 19 dynamics shows that assumptions need to be continually challenged as the science evolves. Examples of how the science has progressed include our understanding of the distinction between influenza and COVID-19 and the dynamics of COVID variants. Models have particular value for decision-making with a novel virus when the dynamics and assumptions are easy for decision-makers to see, and to manipulate. With appropriate adjustment of the parameter values representing transmission risks and infection characteristics, this model could be used to represent any infectious disease. The model could also be adapted for other congregate settings, such as residential facilities.

### Limitations

This model was developed to surface dynamics and thereby aid decision-making about potential mitigation strategies as the pandemic evolves. The model is not intended in its current form to make specific predictions nor to justify specific actions.

Characteristics of households of school community members that influence their exposure to COVID- 19 include recurrent proximity to other household members (number in the household, and overcrowding), intermittent proximity to other individuals who do not live in the household (such as extended family/friends), density of neighborhood housing density (proxy for proximity), and ongoing potential workplace exposures (such as essential or industrial workers in the household. Household behaviors include close physical contact, multiple caregivers of a child, and uses of facial coverings and other safety practices. Household health risks such as presence of individuals with chronic conditions, and/or older age, influences impact of household morbidity from any school- transmitted COVID-19 infection. None of these characteristics are implemented in the current model, for succinctness and due to limited resources for further extending the model, but they may play an important role in local transmission dynamics, particularly since these risk factors often co-occur with one another.

### Future directions

There are three main avenues for further development of this work. First, the model can be extended, adding other agents such as teachers and other school personnel; incorporating more complicated social networks including sibling connections and asymmetric exposures; other interactions such as shared transportation (school buses and carpools) and after-school sports; compliance (reliability) in mitigation such as handwashing and mask-wearing; and more nuanced dynamics for test sensitivity, for example having test sensitivity depend explicitly on symptomatic status rather than only days since infection. Second, the interactions between different input parameters can be explored, by simultaneously varying multiple parameters instead of only one at a time as we have primarily done in this paper. This is a more realistic use of the model and how we envision public health authorities and school systems making use of it. Readers are encouraged to access the model via the user interface at https://agent-based- models.shinyapps.io/RegionalCOVIDSchoolSimulation/ to explore other combinations of input values. Third, the user interface can be augmented by adding side-by-side comparisons of the outcomes for different combinations of input parameter values, narrative descriptions of individual runs of the simulation, and additional outcome time series.

### Conclusion

Models enable stakeholders and modelers to consider infection dynamics and potential mitigation strategies in combination. Public health authorities and school systems can use insights from these models to establish operational needs for safer in-person instruction, such as accurate daily health checks and ongoing timely data on the reliability of mitigation strategies. Models can facilitate an iterative process by which understanding of the system is further deepened, which can in turn be used to reassure communities that schools are capable of delivering in-person instruction without triggering large outbreaks. With future calibration, the model can ultimately have value for prediction.

It is important for decision-making models in COVID-19 to be flexible given the rapid evolution of knowledge about how the virus operates, the rapid transmission dynamics of a disease that spreads through a population exponentially, and the rapidly changing landscape of testing features, costs, and operational burdens. This model can inform decision-making once vaccines are widely available. For example, it provides insight into the potential impact of vaccine exemption decisions and rates.

Public health authorities and school districts can make more meaningful choices about the welfare of K-6 students, their teachers, and their families if these decisions about in-person instruction are based on information from models that incorporate their local conditions and use the different elements available to them, especially those that reflect the COVID-19 situation in the real world. This study provides one such model, recognizing that not all possible elements may be politically or operationally feasible given the characteristics of a particular school community.

## Data Availability

No original data was used in this manuscript.

https://agent-based-models.shinyapps.io/RegionalCOVIDSchoolSimulation/

