## Supporting Information for "Modeling infection dynamics and mitigation strategies to support K-6 in-person instruction during the COVID-19 pandemic"

#### 1. Model structure and dynamics

The model structure and parameters are described in this section; default parameter values and their rationales are provided in the following section. An overview of the model is provided in Table 1.

*Table 1: Overview of the model*

| **Model aspect** | **Description** |
|---|---|
| Agents | Students, parents |
| Agent relationships | Students grouped by class within school<br>Students, parents grouped by household |
| Infection risks | Transmissions in school<br>Transmissions in home<br>Community case rate (exogenous infections) |
| Testing | In response to symptoms or exposure<br>Periodic surveillance testing |
| Safety education | Households with self-reported exposures or symptoms are called to offer information about minimizing exposures, accurately reporting exposures and symptoms. |
| Quarantines | If parents or students report exposures or symptoms, student is quarantined.<br>If infection clusters detected in class, whole class quarantined. |
| Outcomes | % students infected<br>% students infected at school<br># days missed per student<br>% of schools with no transmissions<br>% of schools with no detected infection clusters |

##### 1.1 Time scale and horizon

The model begins on a specified baseline date, at which point a number of agents are instantiated with initial characteristics. The model then changes these characteristics on a day-by-day basis via a repeated sequence of updates that represent attestations, exposures, biological testing, and health safety education outreach efforts. The updates end when the model reaches a pre-specified time horizon, such as the end of a school term or academic year.

#### 1.2 Agents

The model contains two types of simulated individuals ("agents"): students and their associated household adults (two per student). We chose to include these agent types as they form the largest proportion of the school community. The model could be extended to include teachers and other school personnel as additional agents who interact with students and with one other.

The students in the model are each assigned to a particular school and class. The number of schools simulated, the number of classes per school, and the number of students per class are all parameters that can be set via the user interface. In the current model, the schools are statistically independent from each other, and the number of schools is effectively the number of simulation replications used to estimate average outcomes. Simulating more schools results in more precise estimation of the average outcomes for a given scenario.

Another parameter is the number of "close classmates" each student has, representing proximity due to seating arrangement, sharing meals, or other reasons. Close classmates have higher risks of COVID transmission when infectious, compared to the rest of the students in the class. If the "number of close classmates" parameter is set a nonzero value, each class is divided into subgroups of that size; all of the students within each subgroup are "close classmates" with each other. If the "number of close classmates" parameter value is not an integer divisor of the class size, one subgroup will be smaller than the rest.

##### 1.2.1 Initialization of Agent Characteristics

Each run of the model begins at a baseline time point prior to the start of in-person classes. Each agent is assigned initial characteristics with distributions specified by model parameters (see Section 2.3). Initial infection status (actively infected, already recovered from infection, vaccinated, or susceptible to infection) is assigned by sampling without replacement, so that the initial proportions of these statuses among the agents exactly match the corresponding parameter values. For individuals who are actively infected at baseline, the elapsed time since infection at baseline is simulated from a uniform distribution over the length of an infection.

Each individual is also assigned a characteristic representing "COVID safety knowledge" (modeled as a binary variable with options "knowledgeable" and "not knowledgeable"), which affects both their exogenous infection risks and their self-report attestation accuracy (described in greater detail in Section 1.3) over the course of the simulation. Each individual whose "COVID safety knowledge" value is initially "not knowledgeable" is also assigned a binary "receptive to outreach" characteristic; "receptive" individuals will change to the "knowledgeable" state of "COVID safety knowledge" if outreach is made, while "nonreceptive" individuals will remain "not knowledgeable" even if outreach is attempted.

Each individual is also randomly assigned a "symptomatic if infected" characteristic, representing whether that individual will show symptoms if they become infected with COVID.

A baseline test result is generated for each student based on their initial infection status. These results represent a round of school-wide testing prior to school re-opening. The model includes processing time for these tests. The probability of a positive test result is determined by the tested individual's baseline infection characteristics, as well as the sensitivity and specificity of the test.

#### 1.3 Model dynamics

Starting from the baseline date, the simulation performs the following sequence of daily updates to the model.

##### 1.3.1 Infectiousness and symptomatic status

First, the model determines the current symptomatic status and infectious status for each infected individual. Currently, these states are deterministic (non-random) functions of days since infection and the "symptomatic if infected" characteristic; these functions are specified by input parameters (see Section 2.9).

##### 1.3.2 Processing newly returned test results

Next, any newly returned test results are processed. Each specimen takes some time to analyze and return so each test result has a "return date" one or more days after the "collection date". The turnaround time between these dates is set by a parameter specific to the type of test (baseline, surveillance, or positive attestation follow-up). The test result is determined based on the tested individual's characteristics on the collection date, but positive test results do not begin to affect school attendance status until the return date.

When a student tests positive for COVID for the first time, they are quarantined from the test's return date until a parameter-specified amount of time after the collection date. If an adult in a household tests positive for the first time and the student in that household has not yet tested positive, a quarantine is applied to that student. If several students from the same class have tested positive for the first time and been in school within a specified time window, that class is considered to be experiencing an outbreak, and the status of all students in that class changes to quarantine, consistent with outbreak policies of most U.S. public health authorities.

##### 1.3.3 Self-report attestations

Next, each student reports whether or not they or anyone in their household is experiencing COVID symptoms or may have been exposed. The probability of a positive attestation depends on each individual's current infection and symptomatic status. Risk behaviors are not operationalized in the model but are implicitly represented by an increased chance of a positive attestation when an individual is currently infected but in the presymptomatic period of infection. Each student with a positive attestation moves into a quarantine status with a follow-up call scheduled.

##### 1.3.4 Surveillance testing sample collection

The model can include periodic surveillance testing of asymptomatic students, in other words those who are not reporting symptoms or possible exposure. The day(s) of surveillance testing can be specified. If surveillance testing is scheduled for the current day, then a random sample of the students who are in attendance that day (i.e., excluding any currently in quarantine) are selected for surveillance testing. The number of students selected is specified by the "Testing Fraction" parameter. The return date for these tests is determined by the parameter "Lab turnaround time for surveillance test results".

##### 1.3.5 In-school transmission of COVID infection

Next, in-class COVID transmission is simulated. Each currently infectious student who is currently in school has a chance to infect each other student in their class who is not yet infected. The risk of infection for a given student is $1 - (1 - p_C)^C(1 - p_D)^D$, where $C$ is the number of infectious close classmates currently in attendance, $p_C$ is the parameter for the risk of transmission to close classmates per infectious student (the "effective contact risk" for close classmates), $D$ is the number of infectious contacts (including both close and distant classmates) currently in attendance, and $p_D$ is the parameter of risk of transmission to distant classmates per infectious student. For example: if on a given day, a particular student has 2 infectious close classmates and 3 infectious distant contacts currently at school with them, then if $p_C = 0.01$ and $p_D = 0.005$, that student has a $[1 - (1 - 0.01)^2(1 - 0.005)^{2+3}] \times 100\% \approx 4.4\%$ chance of being infected in school on that day.

##### 1.3.6 Outside-school infections

Next, outside-school infections are simulated. Each student has a daily risk of infection outside of school and home, which depends on whether they have COVID-19 safety knowledge. Each household adult also has a risk of exogenous infection.

##### 1.3.7 In-home infections

Next, in-home transmissions are simulated. Infectious students have a chance to infect their household adults, and infectious household adults have a chance to infect their students and a chance to infect the other household adult (if not already infected). For easier interfacing with the available literature, the daily transmission risks are specified indirectly. The user interface provides parameters for the risk of transmission per infection. The risk per day is calculated based on this parameter and the duration-of-infectiousness parameters ("infection time-course"), as:

$$\text{risk per infectious day } = 1 - (1 - \text{risk per infection})^{1/\#\text{ days infectious}}.$$

##### 1.3.8 Outreach to positive attestations

A school district may have an exposure management/contact tracing unit that can respond to families with positive attestations to guide their next actions. At this step of the model, any attestation follow-up calls scheduled for the current day are simulated. The adults and students in the contacted households are all considered to have received outreach about the safety issue that they reported, in these discussions. As described in Section 1.2.1, some individuals have a "receptive to outreach" characteristic; when these participants receive outreach, their COVID safety knowledge characteristic is updated to "knowledgeable", which decreases their daily exogenous infection risks and increases the accuracy of their attestations (parameters "Decrease in exogenous risk from COVID safety education " and "Increase in attestation accuracy after education").

For example, if the parameter "Attestation sensitivity if infected and symptomatic" is set to 60% and the parameter "Increase in attestation accuracy after education" is set to 30%, then a person who is not yet knowledgeable about COVID safety will report symptoms 60% of the time if they have symptoms, whereas a person who is knowledgeable about COVID symptoms (either due to their baseline characteristics or after receiving a follow-up call) will report symptoms 60% + (100% - 60%) × 30% = 72% of the time if they have symptoms. This knowledge variable also has an analogous effect on attestation specificity (the probability of reporting absence of symptoms when they are truly absent).

##### 1.3.9 Follow-up testing of household adults

Finally, any follow-up tests of household adults scheduled for the current day are simulated. The return date for these tests is determined by the parameter "Lab turnaround time for attestation-triggered test results".

#### 1.4 Model outcomes

Note that detected infection clusters are different from actual clusters; actual clusters may remain undetected, and clusters could be mistakenly detected where they do not actually exist, due to false positives. The percentage of schools with no detected infection clusters could change between scenarios either because the rate of actual clusters changes or because the school's ability to detect them changes. Therefore, increases and decreases in this statistic should not be interpreted as "better" or "worse". We included this statistic primarily to contrast it with #4; in many scenarios that we considered, most schools appear to be cluster-free, when in fact on-campus transmission is occurring.

The unit of analysis for calculating outcomes is the school; classes within a school are somewhat correlated, since routine surveillance testing sample selection is conducted at the school level, whereas each school is simulated independently from the others. Future additions to the model, such as transmissions during shared transportation (buses, carpools), after-school sports, recess, lunchtime, and teacher breakroom interactions, may add additional sources of within-school correlation.

### 2. Selection of default parameter values

The default values of all parameters are shown in Table 2. There is uncertainty about the typical clinical time course for coronavirus infection, especially among younger individuals, and about the modifying effects of individuals' baseline characteristics on that time course. Some parameters represent interactions between biological, behavioral, and epidemiological factors; for example, the risk that a person will become infected at a given point in time depends on their biological susceptibility at that moment, their behavior (e.g., social distancing, hand washing), as well as the probability that the people they are interacting with are contagious (and those people's behaviors, e.g., mask wearing). All these factors can vary over time, making it difficult to estimate infection risks precisely.

All parameters are directly controllable through the user interface to enable users to explore plausible scenarios relevant to them. The default parameter values represent a possible set of assumptions. The following section provides our reasoning for some of the default parameter values.

#### 2.1 Size of simulation

##### 2.1.1 Number of schools

Each school in the simulation is simulated independently of the others; therefore, schools can be considered as replicate runs of the simulation (although in our implementation they are all simulated simultaneously, using vectorized code). Therefore, the number of schools is effectively the number of samples drawn from the simulation data-generating distribution, and this number determines the precision of our estimates about this distribution. In this paper, we have chosen to simulate 1000 schools for each scenario; the online user interface defaults to 10 schools, for faster although less precise results.

###### 2.1.2 Weeks of in-person class (time horizon)

We used a time horizon of two months (eight weeks) after start of in-person classes. This time horizon was chosen by balancing competing objectives. Longer time horizons result in larger the cumulative infection rates, making the differences between scenarios easier to distinguish. However, longer time horizons also take longer to simulate. The results presented in Section 3 below took several days to simulate using a distributed computing cluster (the Hoffman2 Shared Cluster provided by UCLA Institute for Digital Research and Education's Research Technology Group).

##### 2.2 School structure

###### 2.2.1 Students per class

The default model uses class size of 15 (parameter "Students per class"), which is used in some school districts and represents about two-thirds of the typical elementary school class size in California.[1]

###### 2.2.2 Classes per school

The default value is 28 classes per school. This default assumes a school with seven grades, each with two classes of 30 students each prior to the pandemic; these classes have now been split in half, resulting in 28 classes of 15 students each.

###### 2.2.3 Number of close classmates

The default number of close classmates is 5 per student (parameter "Number of close classmates per student"), allowing that any given student may be seated near and/or have more contact with a subset of other students in his/her class. We assume that infectious students have an additional probability of transmitting COVID to these close classmates beyond the baseline probability of transmitting COVID to any classmate (see 2.13 below).

#### 2.3 Initial population characteristics

##### 2.3.1 Cumulative incidence at start of simulation

The default cumulative incidence at baseline is set to 4% for both students and adults; this value was chosen based on the cumulative rates of confirmed cases in Los Angeles County in early December.[2,3]

##### 2.3.2 COVID19 prevalence at start of simulation

The default prevalence of active COVID infections at the start of the simulation is 0.6%, for both students and adults (parameters "Baseline COVID prevalence among adults" and parameter "Baseline COVID prevalence among students"). This default value was chosen based on the rates of new confirmed cases in Los Angeles County in early December.[2,3]

##### 2.3.3 Initial prevalence of COVID safety education

The default prevalence of COVID safety education at baseline was set to 10%, based on the assumption that most families could still benefit from more information.

#### 2.4 Baseline testing

##### 2.4.1 Date of baseline test collection and date when in-person classes start

The calendar dates do not have any impact on the simulation; for example, holidays are not modeled. Only the relative timings (i.e., the number of days between baseline testing and start of in-person classes) affect the simulation outcomes. The primary use of calendar dates is to help make the time-series graphs of model outcomes easier to interpret.

#### 2.5 Surveillance testing

The default for an outbreak is three or more students testing positive in the last 14 days, (parameters "Number of recent infections required to declare a classroom outbreak" and "Time window for detecting classroom outbreaks") with default quarantine of 14 days (parameter "Quarantine length after classroom cluster detected"). These values are consistent with LAC DPH criteria for defining and investigating an outbreak.

###### 2.5.1 Testing fraction

The default for surveillance testing fraction of 25% (parameter "Testing fraction”) is similar to protocols for skilled nursing facility (SNF) surveillance.[4] Lab turnaround time for surveillance testing results has default of 2 days (parameter “Lab turnaround time for surveillance test results”).

##### 2.6 Biological test accuracy

By default, we assumed a PCR-type biological test and chose default test accuracy parameter values consistent with published reports of PCR testing.[5] Other testing methods - e.g., antigen testing - could be simulated by appropriate changes to these parameter values. The default specificity of the test, assumed to be PCR, was 99.9%. The default sensitivity on the day of infection and the two subsequent days was assumed 1 – specificity = 0.1%. The default sensitivity on the third day after infection was 50%, and the default for the fourth day was 70%. The default sensitivity for the fifth day was 95% (“peak sensitivity”). The test remains at peak sensitivity for a default of 3 days, and then begins to decline linearly to a default 50% sensitivity at the end of infection. After recovery, the test sensitivity continues to decrease by 10% per day. The full default sensitivity curve is shown in Figure 2.

##### 2.7 Self-report attestation characteristics

Parameters for attestations include the sensitivity if infected and symptomatic or presymptomatic/asymptomatic (parameters “Attestation sensitivity if infected and symptomatic” and “attestation sensitivity if infected and presymptomatic/asymptomatic, based on knowledge of exposure”).

##### 2.8 Education and testing after positive attestations

The default period that a student with a positive attestation is quarantined is 3 days, which allows time for investigation, testing, and results (parameter “Quarantine length after positive attestation”). The default time for a follow-up call is 1 day (parameter “Response time for contacting households after positive attestations”) to investigate the cause. The default increase in attestation sensitivity after a follow-up call is 30% (parameter “Increase in attestation sensitivity after education”). The test turn-around time default is 2 days (parameter “Lab turnaround time for attestation-triggered test results”).

#### 2.9 Infection time-course

Parameters include “Days from infection until infectious” (default 3 days), “Days from infection until no longer infectious” (default 13 days), “Days from infection until symptomatic” (default 5 days), “Days from infection until no longer symptomatic” (default 15 days), and “Days from infection until no longer actively infected".

#### 2.10 Probability of symptoms if infected

By default, 70% of adults and 50% of students will be symptomatic if infected; these defaults were based on available estimates, although there remains substantial uncertainty in the current literature, and these rates may change over time as the virus mutates. [6–10]

#### 2.11 Exogenous Infection Risks

The default infection risk from sources exogenous to our model (sources other than school and household members, such as social activities without adequate protections) is 0.04% for both students and adults. This default was motivated by the rates of new confirmed COVID-19 cases in Los Angeles County in late November and early December. [2,3]

#### 2.12 At-Home Transmission Risks

By default, each household adult who becomes infected has a 40% chance, over the duration of the infection, of transmitting the infection to the other household adult, and a 16% chance of infecting their student. Each student who becomes infected has a 40% chance of infecting each of their household adults.

#### 2.13 At-School Transmission Risks

We chose a relatively high default transmission risk for infectious close classmates (1% per day). This may be an unrealistically pessimistic assumption, especially if mask use and social distancing are strongly enforced and in-person schooldays are shortened so that on-campus mealtimes are unnecessary. Evidence from districts in other states and countries (Ismail et al. 2020) suggests that transmission is rare, especially between students. Nevertheless, we chose a default of 1%, because this value creates the risk of substantial in-school infection rates; as a result, these rates require fewer simulation repetitions in order to be accurately estimated, and the effects of the various interventions on these infection rates are more pronounced.

*Table 2: Default values of simulation parameters*

| Parameter group | Parameter name | Default Value | Source |
|---|---|---|---|
| Size of simulation | How many schools to simulate | 1000 | Determines precision of simulation results |
| | How many weeks of in-person school to simulate (time horizon) | 8 | |
| School structure | Classes per school | 28 | K-6, 60 students per grade, 15 per class |
| | Students per class | 15 | |
| | Number of close classmates per student | 5 | |
| Initial population characteristics | Baseline cumulative incidence of COVID infection among students | 4% | Los Angeles COVID data Dec. 2020[2,3] |
| | Baseline cumulative incidence of COVID infection among adults | 4% | |
| | Baseline prevalence of active COVID infection among students | 0.6% | |
| | Baseline prevalence of active COVID infection among adults | 0.6% | |
| | Baseline proportion of adults vaccinated | 0% | |
| | Baseline proportion of students vaccinated | 0% | |
| | Baseline prevalence of COVID safety education | 10% | |
| | Probability of receptiveness to COVID safety outreach | 50% | |
| Baseline testing | Days between baseline test collection and start of in-person classes | 30 | |
| | Lab turnaround time for baseline test results | 2 | |
| Surveillance testing | Testing fraction (proportion of enrolled students tested per testing day) | 25% | |
| | Testing days | Mondays | |
| | Lab turnaround time for surveillance test results | 2 days | |
| | Quarantine length after student tests positive | 14 days | |
| | Time window for detecting classroom outbreaks | 14 days | |

| | | | |
|---|---|---|---|
| | Number of recent infections required to declare a classroom outbreak | 3 | |
| | Quarantine length after classroom cluster detected | 14 days | |
| Test accuracy | Test specificity | 99.9% | All defaults in "Test accuracy" section are based on PCR test characteristics [5]. |
| | Test sensitivity on day of infection | 0.1% | |
| | Test sensitivity 1 day after infection | 0.1% | |
| | Test sensitivity 2 days after infection | 0.1% | |
| | Test sensitivity 3 days after infection | 50% | |
| | Test sensitivity 4 days after infection | 80% | |
| | Test sensitivity 5+ days after infection ("peak sensitivity") | 95% | |
| | Days of peak sensitivity | 3 | |
| | Test positivity at end of active infection | 50% | |
| | Daily decline in test positivity after recovery | 10% | |
| Self-Reported Symptom/Risk Attestations | Symptom screening sensitivity (probability of accurate self-reporting when infected and symptomatic) | 90% | |
| | Exposure screening sensitivity (probability of accurate self-reporting when infected and presymptomatic/asymptomatic) | 10% | |
| | Attestation specificity (probability of accurately self-reporting when not infected) | 99.9% | |
| | Quarantine length after positive attestation | 3 days | |
| Education and testing after positive attestations | Response time for contacting households after positive attestations | 1 day | |
| | Increase in attestation accuracy after education | 10% | |
| | Decrease in exogenous risk from COVID safety education | 10% | |
| | Response time from attestation follow-up call until test sample collection | 1 day | |
| | Lab turnaround time for attestation-triggered test results | 2 days | |
| | Student quarantine length after household adult tests positive | 14 days | |
| Infection time-course | Days from infection until infectious | 3 days | |
| | Days from infection until no longer infectious | 13 days | |
| | Days from infection until symptomatic | 5 days | |
| | Days from infection until no longer symptomatic | 15 days | |
| | Days from infection until no longer actively infected | 15 days | |

| | | |
|---|---|---|
| Probability of symptoms if infected | for adults | 70% |
| | for students | 50% |
| Exogenous infection risks: | to students | 0.04% |
| | to parents | 0.04% |
| In-home transmission risks (per infection) | from infectious adult in household to student | 16% |
| | from infectious adult in household to partner | 40% |
| | from student to adult in household | 40% |
| At-school transmission risk (per day) | Risk of transmission to close classmates per infectious student | 0.1% |
| | Risk of transmission to distant classmates per infectious student | 0.05% |

### 3. Detailed sensitivity analysis results

#### 3.1 Number of non-social-distanced classmates per student

The number of "close classmates" with whom a student breaks social distancing while at school (e.g., by eating together) also had a substantial impact on transmission rates (*Table 3*).

*Table 3: Average outcomes (with 2.5% and 97.5% percentiles) for 1000 simulated schools, by number of "close contacts" with whom a student breaks social distancing while at school, with other parameters set to default values*

| n close contacts in class | % of enrolled students infected since baseline (cumulative) | % of enrolled students infected from school (cumulative) | # schooldays quarantined per student (cumulative) | % of schools with no on-campus transmissions so far | % of schools with no detected infection clusters so far |
|---|---|---|---|---|---|
| 0 | 4.47 (2.62, 6.43) | 0.03 (0.00, 0.24) | 1.20 (0.92, 1.54) | 88.0 | 98.9 |
| 3 | 4.47 (2.38, 6.67) | 0.04 (0.00, 0.24) | 1.20 (0.91, 1.52) | 85.5 | 99.2 |
| 5 (default) | 4.46 (2.62, 6.67) | 0.05 (0.00, 0.48) | 1.20 (0.93, 1.55) | 82.0 | 98.9 |
| 10 | 4.46 (2.38, 6.67) | 0.07 (0.00, 0.48) | 1.20 (0.91, 1.54) | 75.4 | 99.1 |

#### 3.2 Test sensitivity

Higher peak biological test sensitivity had minimal effects on infection rates (*Table 4*).

*Table 4: Average outcomes (with 2.5% and 97.5% percentiles) for 1000 simulated schools, by peak test sensitivity, with other parameters set to default values*

| Peak test sensitivity (day 5+ after infection) | % of enrolled students infected since | % of enrolled students infected from school (cumulative) | # schooldays quarantined per student (cumulative) | % of schools with no on-campus | % of schools with no detected infection clusters so far |
|---|---|---|---|---|---|

| | baseline (cumulative) | | | transmissions so far | |
|---|---|---|---|---|---|
| 75% | 4.48 (2.86, 6.43) | 0.05 (0.00, 0.24) | 1.22 (0.89, 1.55) | 81.9 | 99.1 |
| 90% | 4.48 (2.62, 6.43) | 0.05 (0.00, 0.48) | 1.21 (0.92, 1.54) | 81.2 | 99.1 |
| 95% (default) | 4.46 (2.62, 6.67) | 0.05 (0.00, 0.48) | 1.20 (0.93, 1.55) | 82.0 | 98.9 |
| 99% | 4.47 (2.62, 6.43) | 0.05 (0.00, 0.24) | 1.20 (0.91, 1.52) | 80.1 | 99.3 |

#### 3.3 Test specificity

Higher test specificity resulted in fewer school days missed and more schools with no transmissions so far (*Table 5*).

*Table 5: Average outcomes (with 2.5% and 97.5% percentiles) for 1000 simulated schools, by COVID test specificity, with other parameters set to default values (except test sensitivity days 0-2, set to 100% - test specificity)*

| test specificity | % of enrolled students infected since baseline (cumulative) | % of enrolled students infected from school (cumulative) | # schooldays quarantined per student (cumulative) | % of schools with no on-campus transmissions so far | % of schools with no detected infection clusters so far |
|---|---|---|---|---|---|
| 85% | 4.52 (2.62, 6.67) | 0.07 (0.00, 0.48) | 5.07 (3.49, 6.88) | 75.6 | 0 |
| 90% | 4.50 (2.62, 6.43) | 0.07 (0.00, 0.48) | 3.50 (2.40, 4.86) | 75.3 | 1.4 |
| 95% | 4.51 (2.62, 6.67) | 0.06 (0.00, 0.48) | 2.16 (1.62, 2.93) | 77.6 | 32.8 |
| 99% | 4.49 (2.62, 6.67) | 0.06 (0.00, 0.48) | 1.36 (1.03, 1.74) | 78.6 | 93.7 |
| 99.9% (default) | 4.46 (2.62, 6.67) | 0.05 (0.00, 0.48) | 1.20 (0.93, 1.55) | 82.0 | 98.9 |
| 100% | 4.52 (2.62, 6.43) | 0.06 (0.00, 0.48) | 1.19 (0.91, 1.50) | 79.8 | 99.6 |

#### 3.4 Interactions between test specificity and test frequency

Varying test specificity between 99% and 100% did not substantially modify the effects of surveillance testing frequency on infection rates (Table 6, Figure 1). However, there were substantial interactions on the outcome scales for schooldays missed and probability of no detected clusters; the combination of frequent testing and lower test specificity produced substantially higher rates of missed days and lower probability of no detected clusters than either of these factors alone.

*Table 6: Test specificity versus frequency of surveillance testing: average outcomes (with 2.5% and 97.5% percentiles for numeric variables) for 1000 simulated schools, with other parameters set to default values.*

| test specificity | surveillance testing days | % of enrolled students infected since baseline (cumulative) | % of enrolled students infected from school (cumulative) | # schooldays quarantined per student (cumulative) | % of schools with no on-campus transmissions so far | % of schools with no detected infection clusters so far |
|---|---|---|---|---|---|---|
| 99% | Once a week (Mondays) | 4.49 (2.62, 6.67) | 0.06 (0.00, 0.48) | 1.36 (1.03, 1.74) | 78.6 | 93.7 |
| | Twice a week (M/Th) | 4.45 (2.62, 6.43) | 0.05 (0.00, 0.48) | 1.54 (1.17, 2.04) | 81.6 | 77.5 |
| | 3x a week (MWF) | 4.49 (2.62, 6.67) | 0.05 (0.00, 0.24) | 1.75 (1.31, 2.41) | 82.6 | 59.1 |
| | Every weekday (M-F) | 4.49 (2.38, 6.43) | 0.05 (0.00, 0.48) | 2.23 (1.57, 3.14) | 80.7 | 22.7 |
| 99.9% | 1x a week (M) *(default)* | 4.46 (2.62, 6.67) | 0.05 (0.00, 0.48) | 1.20 (0.93, 1.55) | 82.0 | 98.9 |
| | 2x a week (M/Th) | 4.49 (2.62, 6.67) | 0.05 (0.00, 0.24) | 1.23 (0.94, 1.58) | 81.7 | 97.5 |
| | 3x a week (MWF) | 4.50 (2.62, 6.43) | 0.05 (0.00, 0.24) | 1.26 (0.96, 1.60) | 80.3 | 95.9 |
| | Every weekday (M-F) | 4.45 (2.62, 6.43) | 0.04 (0.00, 0.24) | 1.29 (0.98, 1.66) | 83.5 | 94.9 |
| 100% | Once a week (Mondays) | 4.52 (2.62, 6.43) | 0.06 (0.00, 0.48) | 1.19 (0.91, 1.50) | 79.8 | 99.6 |
| | Twice a week (M/Th) | 4.49 (2.86, 6.43) | 0.05 (0.00, 0.24) | 1.20 (0.93, 1.52) | 82.5 | 98.1 |
| | 3x a week (MWF) | 4.46 (2.62, 6.43) | 0.04 (0.00, 0.24) | 1.21 (0.91, 1.54) | 83.7 | 98.7 |
| | Every weekday (M-F) | 4.49 (2.62, 6.43) | 0.05 (0.00, 0.24) | 1.22 (0.92, 1.56) | 83.7 | 97.8 |

*Figure 1: Average outcomes for 1000 simulated schools, by surveillance testing frequency and test specificity, with other parameters set to default values*

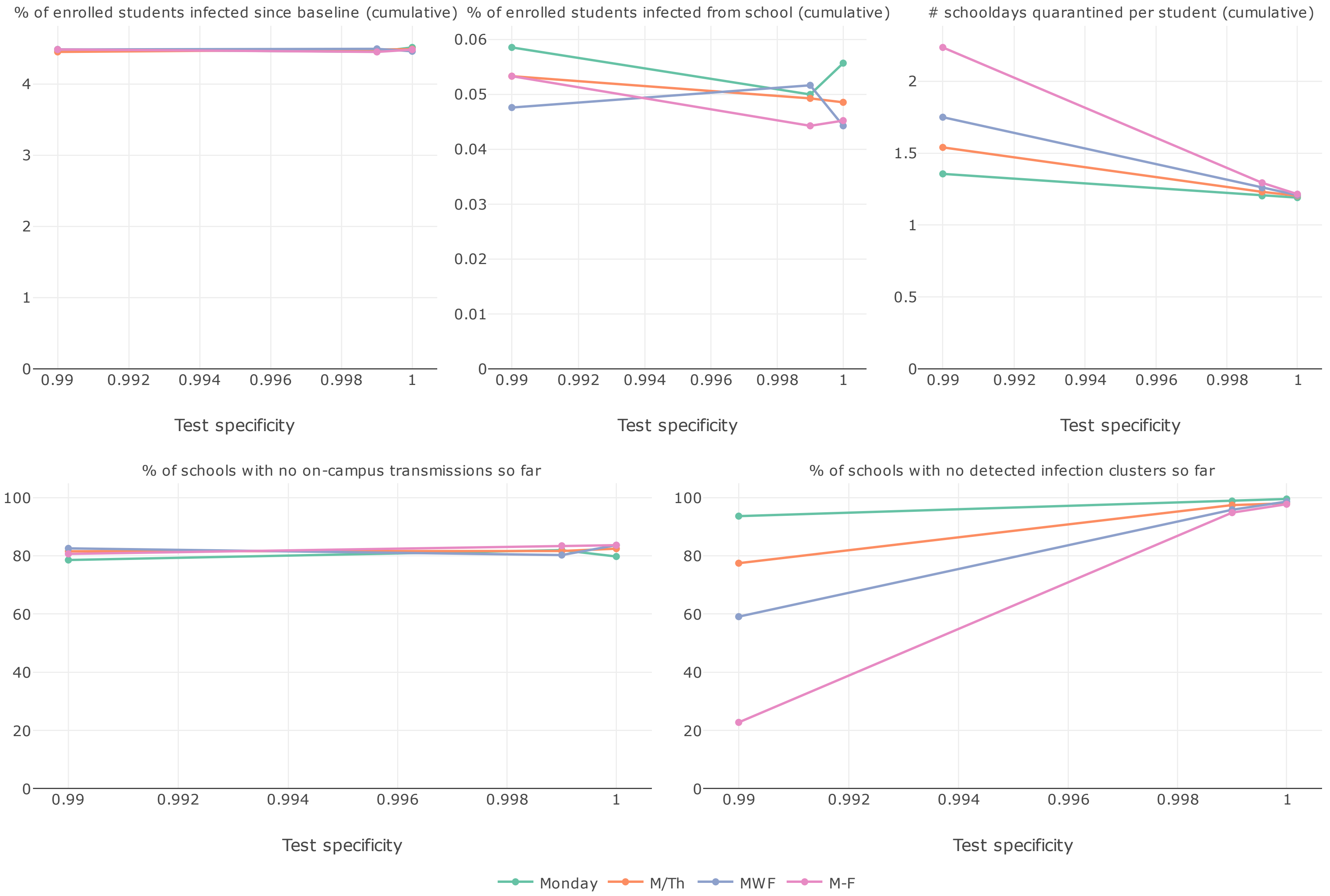

#### 3.5 Risk of transmission to close classmates

The magnitude of the transmission rate between "close contacts" who break social distancing also had substantial impacts on the infection rates (Table 7).

*Table 7: Average outcomes by risk of transmission to close classmates*

| risk per infected close contact | % of enrolled students infected since baseline (cumulative) | % of enrolled students infected from school (cumulative) | # schooldays quarantined per student (cumulative) | % of schools with no on-campus transmissions so far | % of schools with no detected infection clusters so far |
|---|---|---|---|---|---|
| 0% *(no risk)* | 4.47 (2.62, 6.43) | 0.03 (0.00, 0.24) | 1.20 (0.92, 1.54) | 88.0 | 98.9 |
| 0.01% | 4.46 (2.62, 6.43) | 0.04 (0.00, 0.24) | 1.21 (0.92, 1.56) | 86.4 | 98.7 |
| 0.05% | 4.47 (2.62, 6.43) | 0.04 (0.00, 0.24) | 1.21 (0.91, 1.55) | 85.0 | 99.0 |
| 0.10% *(default)* | 4.46 (2.62, 6.67) | 0.05 (0.00, 0.48) | 1.20 (0.93, 1.55) | 82.0 | 98.9 |
| 0.50% | 4.53 (2.61, 6.67) | 0.12 (0.00, 0.71) | 1.21 (0.91, 1.55) | 62.2 | 98.8 |
| 1.00% | 4.62 (2.62, 6.67) | 0.21 (0.00, 0.95) | 1.22 (0.92, 1.58) | 46.6 | 96.8 |

#### 3.6 Risk of transmission to distanced classmates

The magnitude of the transmission rate from infectious students to classmates who maintain distancing had substantial effects on infection rates and also on attendance rates (Table 8).

*Table 8: Average outcomes by risk of transmission to distanced classmates*

| risk per infected distant contact | % of enrolled students infected since baseline (cumulative) | % of enrolled students infected from school (cumulative) | # schooldays quarantined per student (cumulative) | % of schools with no on-campus transmissions so far | % of schools with no detected infection clusters so far |
|---|---|---|---|---|---|
| 0% *(no risk)* | 4.43 (2.62, 6.67) | 0.02 (0.00, 0.24) | 1.20 (0.88, 1.54) | 91.4 | 98.8 |
| 0.01% | 4.44 (2.62, 6.67) | 0.03 (0.00, 0.24) | 1.20 (0.92, 1.52) | 88.2 | 99.4 |
| 0.05% *(default)* | 4.46 (2.62, 6.67) | 0.05 (0.00, 0.48) | 1.20 (0.93, 1.55) | 82.0 | 98.9 |
| 0.10% | 4.48 (2.38, 6.67) | 0.08 (0.00, 0.48) | 1.20 (0.91, 1.54) | 73.5 | 98.6 |
| 0.50% | 4.80 (2.62, 7.38) | 0.40 (0.00, 1.43) | 1.25 (0.93, 1.66) | 26.4 | 91.5 |
| 1.00% | 5.25 (2.86, 8.10) | 0.86 (0.00, 2.62) | 1.32 (0.95, 1.86) | 11.5 | 75.7 |

#### 3.7 Exogenous infection risk

Unsurprisingly, higher exogenous risks resulted in higher rates of infections and school days missed (Table 9).

*Table 9: Average outcomes by exogenous risk*

| exogenous infection risk per day (students and household adults) | % of enrolled students infected since baseline (cumulative) | % of enrolled students infected from school (cumulative) | # schooldays quarantined per student (cumulative) | % of schools with no on-campus transmissions so far | % of schools with no detected infection clusters so far |
|---|---|---|---|---|---|
| 0.004% | 0.58 (0.00, 1.43) | 0.01 (0.00, 0.01) | 0.55 (0.41, 0.71) | 97.5 | 100 |
| 0.040% (default) | 4.46 (2.62, 6.67) | 0.05 (0.00, 0.48) | 1.20 (0.93, 1.55) | 82.0 | 98.9 |
| 0.400% | 34.54 (30.24, 39.29) | 0.27 (0.00, 0.95) | 6.14 (5.29, 7.11) | 33.8 | 14.9 |

#### 3.8 Attestation sensitivity if symptomatic

Higher symptom attestation sensitivity resulted in lower infection rates and slightly higher rates of days missed (Table 10).

*Table 10: Average outcomes by sensitivity of attestations when symptomatic*

| attestation sensitivity when symptomatic | % of enrolled students infected since baseline (cumulative) | % of enrolled students infected from school (cumulative) | # schooldays quarantined per student (cumulative) | % of schools with no on-campus transmissions so far | % of schools with no detected infection clusters so far |
|---|---|---|---|---|---|
| 0% | 4.54 (2.62, 6.91) | 0.10 (0.00, 0.48) | 0.94 (0.70, 1.20) | 65.9 | 98.8 |
| 50% | 4.49 (2.62, 6.43) | 0.05 (0.00, 0.24) | 1.21 (0.92, 1.54) | 80.6 | 98.6 |
| 80% | 4.48 (2.62, 6.43) | 0.05 (0.00, 0.48) | 1.21 (0.92, 1.52) | 80.7 | 98.7 |
| 90% (default) | 4.46 (2.62, 6.67) | 0.05 (0.00, 0.48) | 1.20 (0.93, 1.55) | 82.0 | 98.9 |
| 100% | 4.47 (2.62, 6.67) | 0.05 (0.00, 0.24) | 1.20 (0.91, 1.51) | 81.0 | 98.6 |

#### 3.9 Attestation sensitivity presymptomatic/asymptomatic

Higher attestation sensitivity in the presymptomatic/asymptomatic phase (e.g., higher probability of positive attestations due to suspected recent exposure) result in substantially lower infection rates and slightly higher days missed (Table 11).

*Table 11: Average outcomes by sensitivity of attestations when presymptomatic or asymptomatic*

| attestation sensitivity when presymptomatic/ asymptomatic | % of enrolled students infected since baseline (cumulative) | % of enrolled students infected from school (cumulative) | # schooldays quarantined per student (cumulative) | % of schools with no on-campus transmissions so far | % of schools with no detected infection clusters so far |
|---|---|---|---|---|---|
| 1% | 4.51 (2.62, 6.67) | 0.08 (0.00, 0.48) | 1.15 (0.88, 1.47) | 72.8 | 98.3 |
| 10% (default) | 4.46 (2.62, 6.67) | 0.05 (0.00, 0.48) | 1.20 (0.93, 1.55) | 82.0 | 98.9 |
| 25% | 4.44 (2.62, 6.43) | 0.03 (0.00, 0.24) | 1.27 (0.98, 1.60) | 89.9 | 99.1 |
| 50% | 4.45 (2.62, 6.43) | 0.01 (0.00, 0.24) | 1.32 (1.00, 1.68) | 96.4 | 98.7 |

#### 3.10 Attestation specificity

Higher attestation specificity resulted in higher infection rates (presumably due to effectively increased class sizes) and lower rates of schooldays missed (Table 12).

*Table 12: Average outcomes by specificity of attestations*

| attestation specificity | % of enrolled students infected since baseline (cumulative) | % of enrolled students infected from school (cumulative) | # schooldays quarantined per student (cumulative) | % of schools with no on-campus transmissions so far | % of schools with no detected infection clusters so far |
|---|---|---|---|---|---|
| 95.0% | 4.26 (2.38, 6.19) | 0.01 (0.00, 0.24) | 17.39 (16.76, 18.01) | 94.6 | 98.9 |
| 99.0% | 4.37 (2.62, 6.43) | 0.04 (0.00, 0.24) | 4.90 (4.47, 5.34) | 85.8 | 98.6 |
| 99.9% (default) | 4.46 (2.62, 6.67) | 0.05 (0.00, 0.48) | 1.20 (0.93, 1.55) | 82.0 | 98.9 |
| 100.0% | 4.45 (2.62, 6.67) | 0.06 (0.00, 0.48) | 0.75 (0.48, 1.06) | 77.8 | 98.9 |

#### 3.11 Effect of COVID safety education on self-report attestation accuracy

Increases in attestation accuracy from COVID safety education did not have substantial effects on the outcomes considered here (Table 13).

*Table 13: Outcomes by effect of education on attestation accuracy, with other parameters set to defaults*

| increase in accuracy after education | % of enrolled students infected since baseline (cumulative) | % of enrolled students infected from school (cumulative) | # schooldays quarantined per student (cumulative) | % of schools with no on-campus transmissions so far | % of schools with no detected infection clusters so far |
|---|---|---|---|---|---|
| 0% | 4.48 (2.62, 6.67) | 0.05 (0.00, 0.48) | 1.21 (0.92, 1.51) | 81.5 | 99.2 |
| 10% (default) | 4.46 (2.62, 6.67) | 0.05 (0.00, 0.48) | 1.20 (0.93, 1.55) | 82.0 | 98.9 |
| 25% | 4.49 (2.38, 6.67) | 0.05 (0.00, 0.24) | 1.20 (0.90, 1.53) | 82.6 | 99.0 |
| 50% | 4.46 (2.61, 6.43) | 0.05 (0.00, 0.24) | 1.19 (0.90, 1.51) | 82.2 | 99.3 |

#### 3.12 Effect of COVID safety education on exogenous risk

Larger decreases in exogenous risk from COVID safety education resulted in slightly lower overall infection rates and rates of schooldays missed (Table 14).

*Table 14: Outcomes by effect of education on exogenous risk, with other parameters set to defaults*

| Decrease in exogenous risk after education | % of enrolled students infected since baseline (cumulative) | % of enrolled students infected from school (cumulative) | # schooldays quarantined per student (cumulative) | % of schools with no on-campus transmissions so far | % of schools with no detected infection clusters so far |
|---|---|---|---|---|---|
| 0% | 4.54 (2.62, 6.67) | 0.05 (0.00, 0.24) | 1.22 (0.93, 1.55) | 80.4 | 98.8 |
| 10% (default) | 4.46 (2.62, 6.67) | 0.05 (0.00, 0.48) | 1.20 (0.93, 1.55) | 82.0 | 98.9 |

| | | | | | |
|---|---|---|---|---|---|
| 25% | 4.37 (2.62, 6.43) | 0.05 (0.00, 0.48) | 1.19 (0.89, 1.51) | 81.5 | 98.8 |
| 50% | 4.18 (2.38, 6.19) | 0.05 (0.00, 0.48) | 1.16 (0.88, 1.52) | 81.2 | 98.6 |

#### 3.13 Probability of receptiveness to COVID safety education outreach

Higher probabilities of receptiveness to COVID safety education outreach resulted in slightly lower rates of infections and schooldays missed (Table 15).

*Table 15: Outcomes by probability of receptivity to COVID safety education outreach, with other parameters set to defaults*

| Probability of receptiveness to education outreach | % of enrolled students infected since baseline (cumulative) | % of enrolled students infected from school (cumulative) | # schooldays quarantined per student (cumulative) | % of schools with no on-campus transmissions so far | % of schools with no detected infection clusters so far |
|---|---|---|---|---|---|
| 0% | 4.54 (2.62, 6.67) | 0.06 (0.00, 0.48) | 1.21 (0.92, 1.55) | 77.5 | 98.9 |
| 50% (default) | 4.46 (2.62, 6.67) | 0.05 (0.00, 0.48) | 1.20 (0.93, 1.55) | 82.0 | 98.9 |
| 100% | 4.44 (2.62, 6.43) | 0.05 (0.00, 0.48) | 1.20 (0.92, 1.55) | 81.1 | 99.4 |

#### 3.14 Surveillance testing fraction (additional scenarios)

*Table 16: Average outcomes (with 2.5% and 97.5% percentiles) for 1000 simulated schools, by surveillance testing fraction (% enrolled students tested per testing day), with other parameters set to default values*

| Testing fraction | % of enrolled students infected since baseline (cumulative) | % of enrolled students infected from school (cumulative) | # schooldays quarantined per student (cumulative) | % of schools with no on-campus transmission s so far | % of schools with no detected infection clusters so far |
|---|---|---|---|---|---|
| 0% (No surveillance) | 4.48 (2.62, 6.67) | 0.05 (0.00, 0.48) | 1.18 (0.89, 1.51) | 81.4 | 99.5 |
| 10% | 4.51 (2.62, 6.67) | 0.05 (0.00, 0.48) | 1.20 (0.90, 1.52) | 80.5 | 99.4 |
| 25% (*default*) | 4.46 (2.62, 6.67) | 0.05 (0.00, 0.48) | 1.20 (0.93, 1.55) | 82.0 | 98.9 |
| 50% | 4.44 (2.62, 6.67) | 0.05 (0.00, 0.24) | 1.23 (0.93, 1.56) | 81.8 | 97.6 |
| 100% | 4.42 (2.86, 6.20) | 0.04 (0.00, 0.24) | 1.26 (0.98, 1.63) | 84.5 | 95.5 |

*Figure 2: Default sensitivity (probability of positive result) for biological testing in the simulation model, by infection duration.*

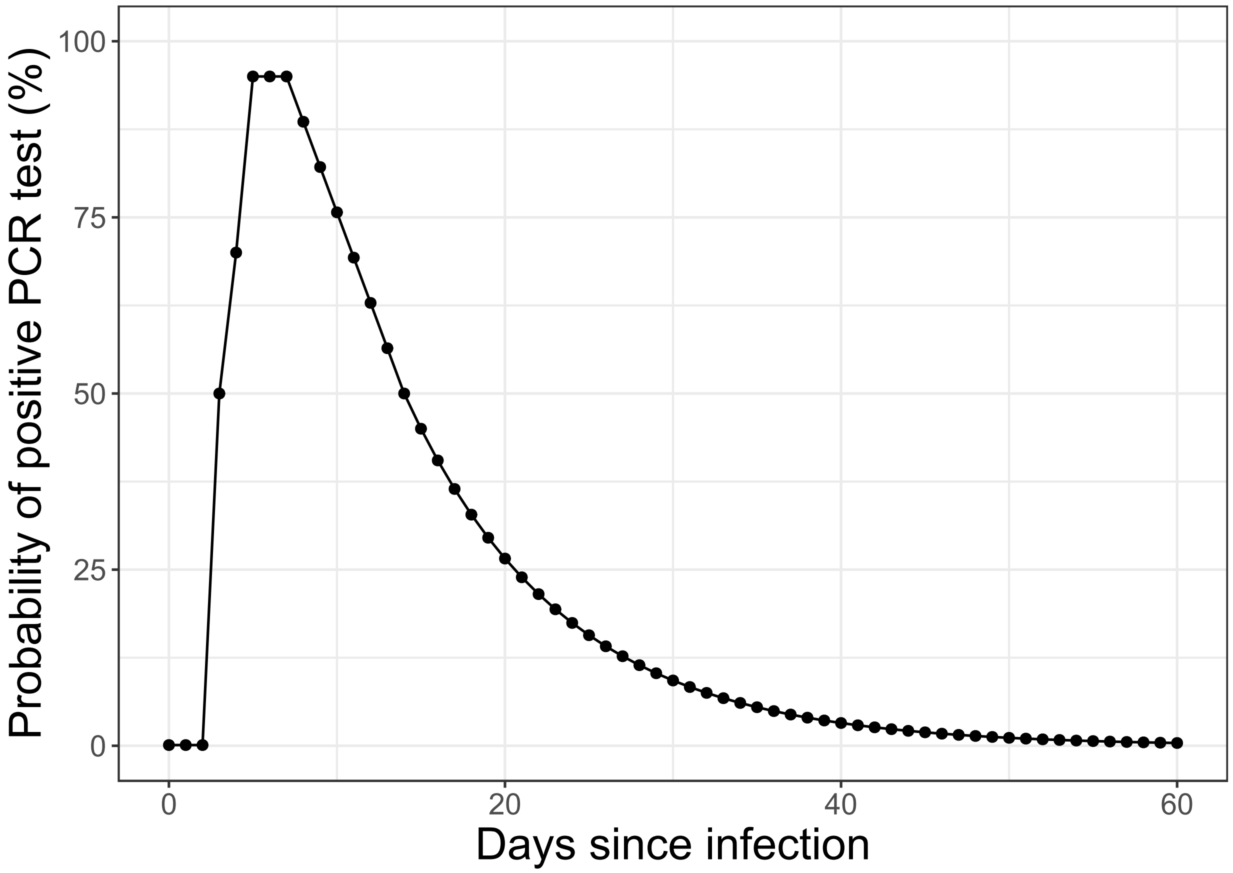


#### 4. Tornado plots

Figure 3 - Figure 7 show tornado plots summarizing the ranges of values for each outcome in the sensitivity analyses above.

*Figure 3: Ranges of average rates of enrolled students infected since baseline, varying one variable at a time with other variables at default values (vertical line indicates default scenario)*

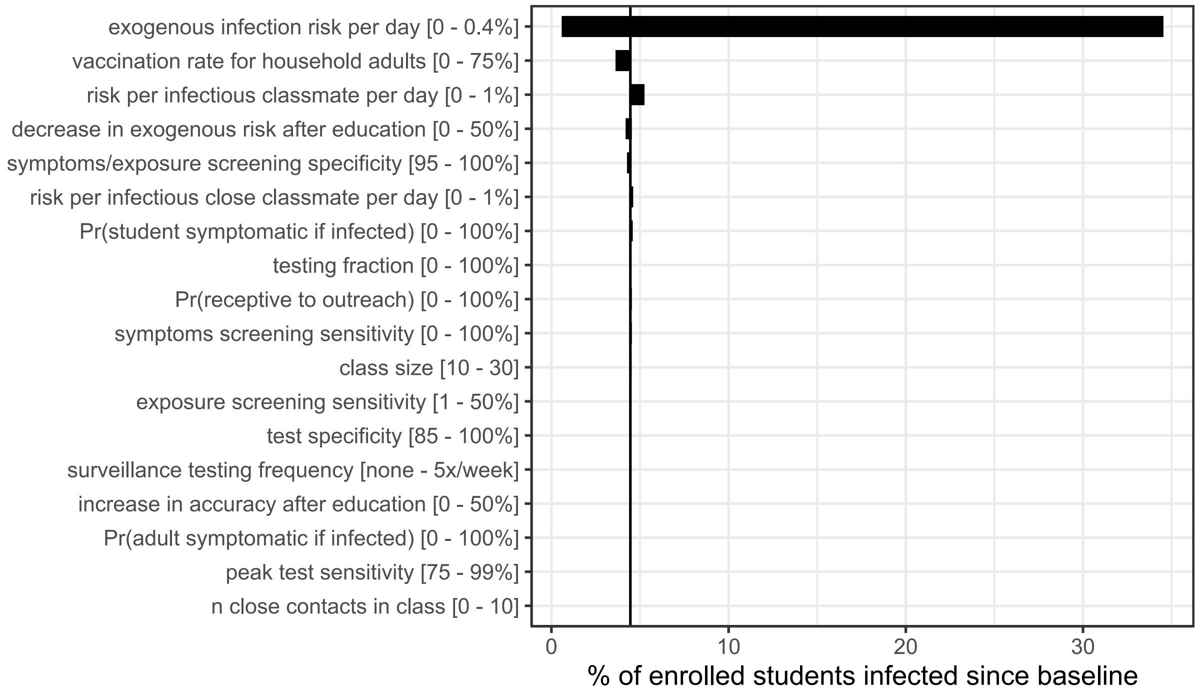

*Figure 4: Ranges of average rates of enrolled students infected from school, varying one variable at a time with other variables at default values (vertical line indicates default scenario)*

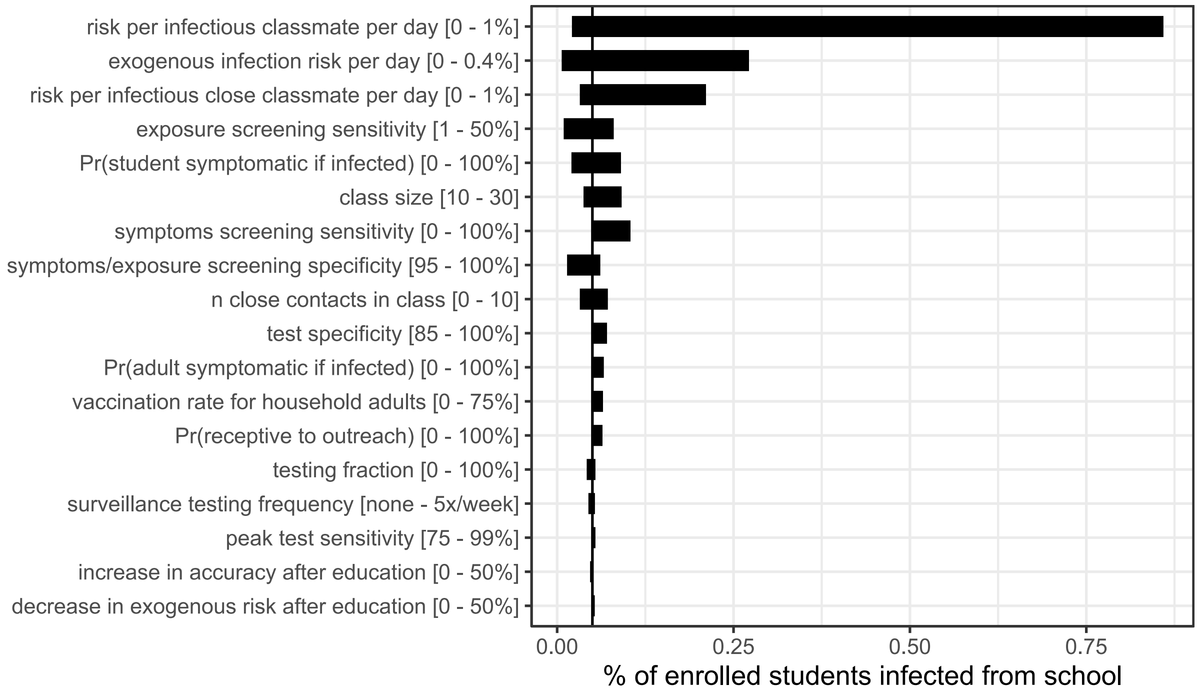

*Figure 5: Ranges of average rates of schooldays missed, varying one variable at a time with other variables at default values (vertical line indicates default scenario)*

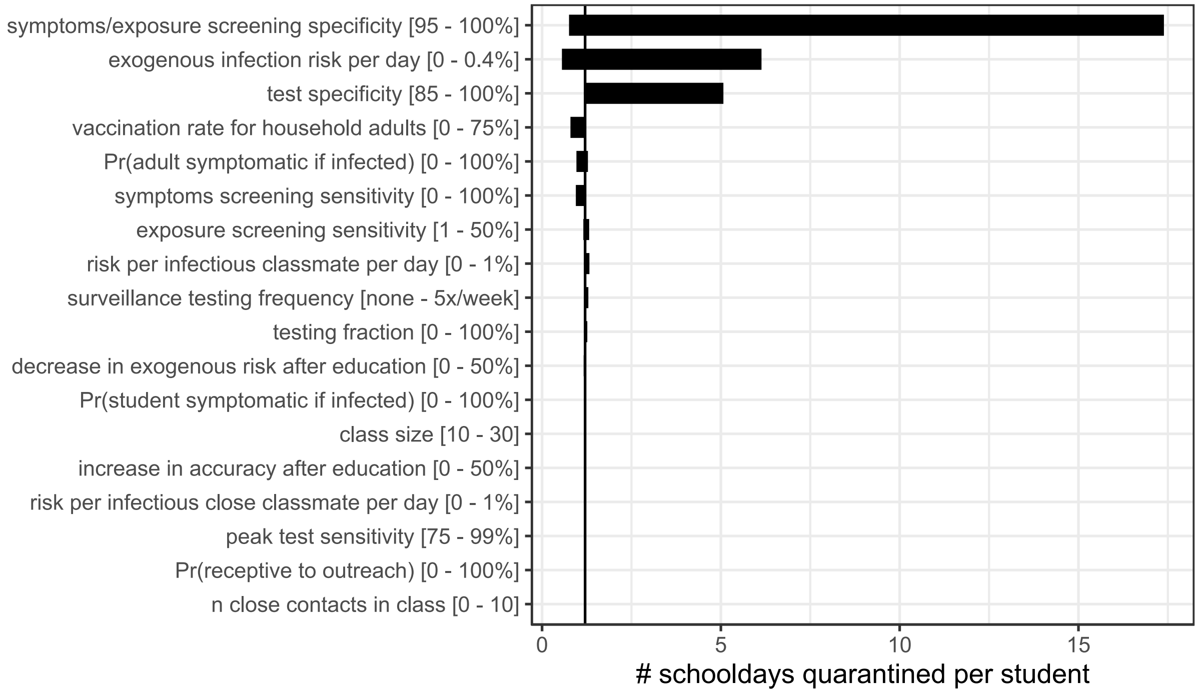

*Figure 6: Ranges of % of schools with no on-campus transmissions, varying one variable at a time with other variables at default values (vertical line indicates default scenario)*

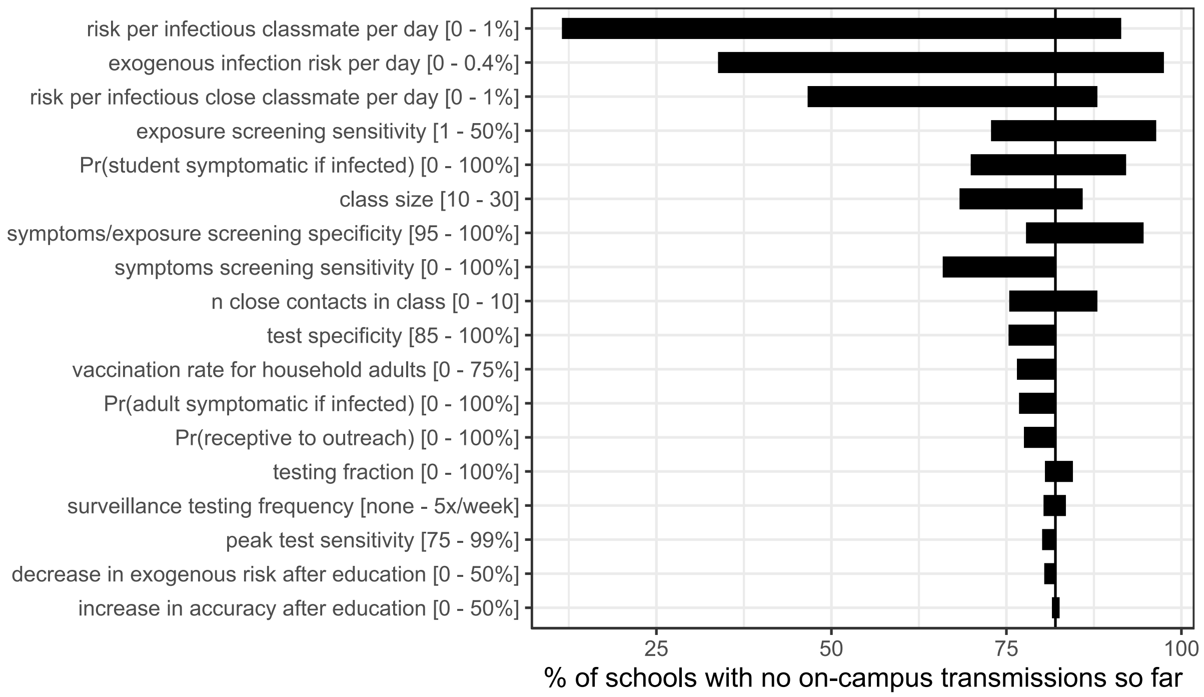

*Figure 7 Ranges of % of schools with no detected infection clusters, varying one variable at a time with other variables at default values (vertical line indicates default scenario)*

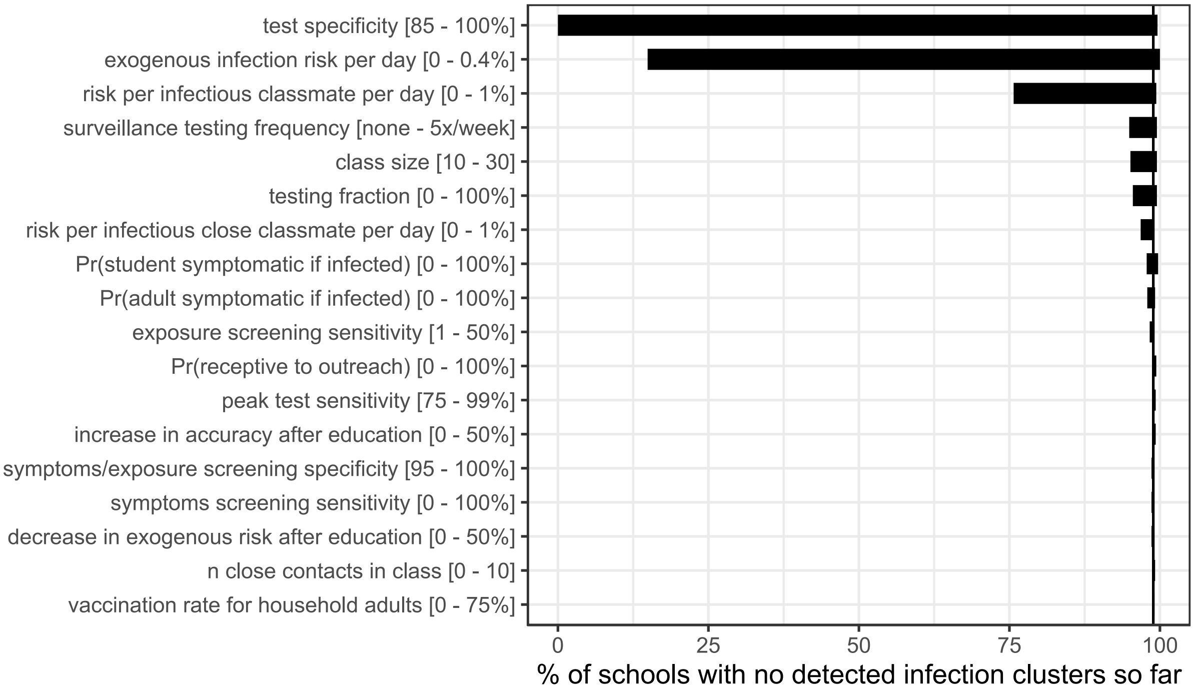
